# Epidemiological overlaps in COVID-19 and malaria within healthcare and community settings of Southern Ghana

**DOI:** 10.1101/2023.12.04.23299372

**Authors:** Gloria Amegatcher, Maame E. Acquah, Deborah Tetteh, Rachael Obeng, Ethel Debrah, Bridget Quist, Priscilla Acquah-Jackson, Kyerewaa A. Boateng, Gideon Twieku, Samuel Armoo, Gordon A. Awandare, Lydia Mosi, Charles A. Narh

## Abstract

**Background:** COVID-19 disruptions in Africa in 2020-2022 contributed to reductions in malaria control activities including antimalarial surveillance programs. This study investigated the malaria burden and distribution on the background of active transmission of SARS-CoV-2 in Southern Ghana. Specifically, it aimed to identify epidemiological factors that can maximise programmatic control for both diseases, utilising community health education and medical screening (CHEMS).

**Methods:** Between October-December 2022, prospective cross-sectional surveys, with CHEMS were conducted in Greater Accra and Central regions, where 994 participants enrolled either at a hospital or community setting provided demographic and clinical data including history of clinical malaria infection and antimalarial treatment in the past two weeks. Of this study population, 953 provided nasal/throat swabs for COVID-19 RT-PCR testing, with a subset of 136 participants also providing finger-prick blood for malaria RDT testing.

**Results:** The study population comprised of 73.6% adults, with 54.6% COVID-19 vaccination rate. Overall, 18.1% of participants had a history of clinical malaria, which was associated (adjusted odds ratio > 1.50, P-value ≤ 0.022) with COVID-19 symptoms and positivity, study area and hospital setting, suggestive of overlaps in the epidemiological risk for malaria. On a background of widespread SARS-CoV-2 infections (12-37%), malaria parasitaemia was detected in 6%, with 2% being co-infections. Among the malaria positives, 9.5% had a history of antimalarial treatment, which suggested that their infections were recrudescent parasitaemia.

**Conclusion:** The overlaps in the epidemiological risk for malaria and COVID-19 indicate that innovative surveillance programs, with community engagement are needed to maximise control interventions including treatment of asymptomatic malaria infections.

## Introduction

COVID-19 disruptions accounted for 14 million increase in malaria cases in the sub-Sahara Africa (SSA) region in 2020 ^1^, indicating that innovative control programs are needed to supplement the activities of National Malaria Control/Elimination Programs (NMCPs or NMEPs). Malaria remains a major public health threat in SSA, with children under five years and pregnant women being the most vulnerable ^1^. Since 2020, due to resource reallocation to and prioritisation of COVID-19 activities, there have been significant reductions in malaria control and surveillance activities including diagnosis, treatment and antimalarial resistance monitoring ^1,2.^

Malaria surveillance in Ghana, mostly focused on clinical infections, was disrupted by the COVID-19 pandemic. For instance, there were increased cases of malaria after the first wave of COVID-19 in March 2020 ^3^, with low treatment-seeking at health facilities ^4^. Public engagement in infectious diseases surveillance in Ghana remains a challenge for control programs ^5^. Community Health Education and Medical Screening (CHEMS), an incentivised approach to community participation and engagement in public health, implemented by the University of Ghana to sensitise communities to COVID-19 interventions ^6^, can provide synergy for malaria surveillance programs at the national level.

Indeed, significant overlaps in the clinical presentations and epidemiological distribution of malaria, caused by *Plasmodium falciparum*, and COVID-19 cases in endemic communities in Ghana have contributed to malaria misdiagnosis and impacted the effectiveness of interventions ^7^. While uptake of interventions at the community level declined during the COVID-19 disruptions ^8^, there is limited data to assess malaria case and co-infection rates in communities with active SARS-CoV-2 transmission.

Our group was actively involved in several national COVID-19 response initiatives between 2020-2023, leveraging programs including CHEMS to diagnose and track SARS-CoV-2 transmission networks ^6,9,10^. Therefore, amidst the limited resources and disruptions in malaria control programs, we tested the feasibility of integrating malaria surveillance into these COVID-19 initiatives, with the aim of determining the distribution of the disease and its epidemiological overlaps with COVID-19 in southern Ghana.

## Methods

This study was embedded within a COVID-19 pre-clinical study that aimed to assess the clinical performance of a novel point-of-care SARS-CoV-2 RNA test. The details of the pre-clinical study have been described elsewhere (Trial Registry # ACTRN12623000066684). Briefly, between October-December 2022 (Figure 1A), 1,065 participants were recruited through multi-site prospective cross-sectional surveys in eight communities in Greater Accra and four communities in the Central regions of Ghana, with the CHEMS approach being pivotal to community acceptance and engagement in the study.

**Figure 1:**
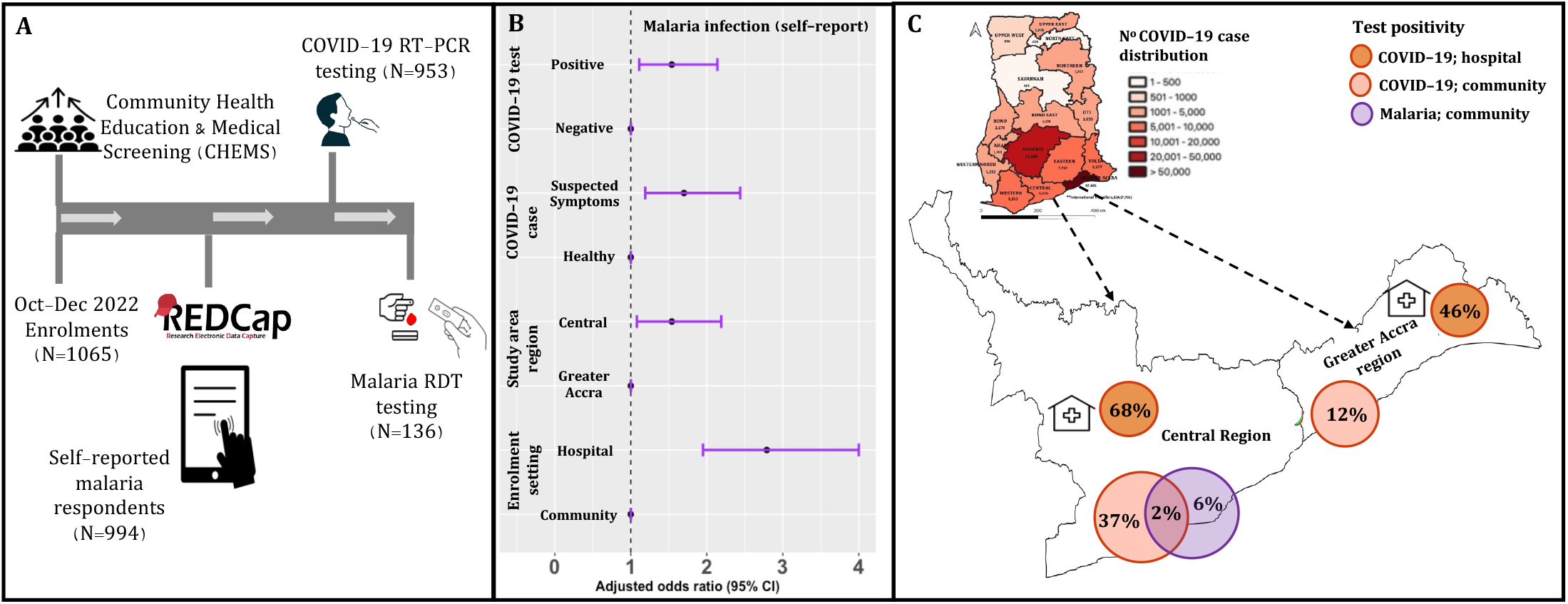
Enrolment with COVID-19 and malaria testing in the Greater Accra and Central regions of Ghana. **A**. Enrolment, CHEMS and testing for SARS-CoV-2 (both study areas) using throat and/or nasal swabs, and malaria RDT testing using finger-prick blood (only Central region). **B**. The adjusted odds ratio for reported malaria in the past two weeks. The reference variables are indicated with odds ratio of 1. **C**. The map of Ghana shows cumulative COVID-19 case distribution in all 16 regions based on routine surveillance data made publicly available by the Ghana Health Services ^12^. The extended map shows the COVID-19 and malaria infection rates for the study areas: Greater Accra and Central regions. The malaria infection rate for this study was assessed only in the study communities in the Central region.

In each community, the participants were recruited through convenience sampling at hospitals/clinics (hospital setting) and at public places (community setting) including markets and schools (Figure 1A). Through CHEMS, the field staff educated the community and raised awareness about COVID-19 testing and vaccination, malaria diagnosis and treatment, and clinical symptoms of both diseases.

Additionally, participants were provided with free medical screening including body temperature and weight checks and free private consultation with the study nurse. Eligible participants provided nasal/throat swabs for COVID-19 RT-PCR testing (2019-nCoV kit, Sansure Biotech and Allplex™ SARS-CoV-2 Master Assay, Seegene), with optional finger-prick blood for malaria RDT testing (Bioline™ Malaria Ag P.f test, Abbot) (Figure 1A). The test results of the novel point-of-care test were used to confirm SARS-CoV-2 infection in samples with invalid RT-PCR test results. The participants’ demographic and clinical data including self-reported malaria infection and antimalarial treatment in the past two weeks were recorded in REDCap. Where available, positive cases (malaria or COVID-19) were referred to the nearest hospital/clinic.

For this study, participants were categorised into age groups: < 18 (children), 18-59 (adults) and ≥ 60 (elderly); Statistical analyses including comparison of proportions and logistic regression were performed as described elsewhere ^11^ using R v3.5.2 and STATA SE 18. Age, gender and catchment area were considered potential confounders and were adjusted for in the final regression model. The adjusted odds ratios with 95% confidence interval (CI) were plotted (Figure 1B) with the ggplot2 R package. Statistical significance; P-value < 0.05.

## Results

Of the 1,065 eligible participants enrolled, 994 responded to questions on malaria infection, and therefore they constituted the final study population. Of this, 953 provided swabs for COVID-19 testing and 136 provide finger-prick blood for malaria testing (Figure 1A).

The demographics of the study population is shown in Table 1, focusing on self-reported malaria infection stratified by the study variables. The majority of the study population comprised of adults (73.6%) and females (61.1%). Based on COVID-19 case definition ^13^, 74.9% of the study participants reported having no clinical symptoms associated with COVID-19 (hereafter referred to as “healthy participants”) and 23.1% reported having COVID-like symptoms (“suspected cases”). COVID-19 case confirmation by RT-PCR testing resulted in 39% positivity in the study population. At the community and hospital settings, 12% and 46%, respectively, were recorded for Greater Accra, and 37% and 68%, respectively, for the Central region (Figure 1C). The COVID-19 vaccination rate overall was 54.5%, and 91.6% of the participants reported not having COVID-19 in the last one month.

**Table 1:** Malaria infection in the study population in the Central region of Ghana.

| Demographic characteristics |  | Self-reported malaria (N=994) |  |  | Malaria RDT testing (N=136) |  |  |
| --- | --- | --- | --- | --- | --- | --- | --- |
|  |  | Total (N) | YES (%) | P-value | Total (N) | Positivity (%) | P-value |
| Age groups | Children | 197 | 17.8 | 0.988 | 82 | 2.4 | 0.102 |
|  | Adults | 732 | 18.2 |  | 47 | 10.6 |  |
|  | Elderly | 65 | 18.5 |  | 7 | 14.3 |  |
| Gender | Male | 384 | 20.3 | 0.187 | 42 | 4.8 | 0.700 |
|  | Female | 602 | 16.8 |  | 93 | 6.5 |  |
| Catchment area<br>(Region) | Greater Accra | 365 | 14.2 | 0.020 | 0 | 0.0 | NA |
|  | Central | 629 | 20.3 |  | 136 | 5.9 |  |
| Enrolment setting | Community | 619 | 12.9 | 0.001 | 136 | 5.9 | NA |
|  | Hospital/Clinic | 375 | 26.7 |  | 0 | 0.0 |  |
| COVID-19 vaccination | Vaccinated | 531 | 16.4 | 0.105 | 40 | 7.5 | 0.429 |
|  | Unvaccinated | 441 | 20.6 |  | 93 | 4.3 |  |
| Had COVID-19 past 1-month | YES | 80 | 17.5 | 0.920 | 24 | 4.2 | 0.680 |
|  | NO | 874 | 18.0 |  | 110 | 6.4 |  |
| COVID-19 case | Suspected | 231 | 25.5 | 0.003 | 17 | 0.0 | 0.595 |
|  | Healthy | 688 | 16.4 |  | 118 | 6.8 |  |
| Confirmed COVID-19 | Positive | 373 | 22.8 | 0.022 | 54 | 1.9 | 0.211 |
|  | Negative | 580 | 15.0 |  | 82 | 8.5 |  |
| Malaria treatment | Clinical treatment | 125 | 99.2 | 0.520 | 21 | 9.5 | 1.000 |
|  | Self-medication* | 52 | 100.0 |  | 5 | 100.0 |  |

Not applicable (NA) as malaria RDT testing was only completed in the community setting in the Central region only. \*Self-medication includes participants who reported non-antimalarial treatment and no treatment. #Clinical treatment includes participants who reported being given approved antimalarials at the hospital or pharmacy.

Next, we determined the association between malaria infection, in the previous two weeks, and epidemiological factors including COVID-19 disease. Overall, 18.1% of participants reported having clinical malaria (Table 1), of which 70.6% received clinical treatment (antimalarials from the hospital or pharmacy). Self-medication included the use of non-antimalarials that were not clinically prescribed. Self-reported clinical malaria was significantly associated (adjusted odds ratio > 1.50, P-value ≤ 0.022) with COVID-19 (suspected symptoms and test positivity), enrolment setting (hospital/clinic) and study area (Central region) (Figure 1B).

We then conducted malaria RDT testing in the Central region, which had the highest risk of self-reported malaria. Confirmation of *P. falciparum* infections among the subset of 136 participants (60.3% being children) resulted in a parasite prevalence of 6.0%; The majority being adults with no history of clinically treated malaria in the previous two weeks, suggesting that the current infections were recently acquired. A minority, 9.5%, had history of clinically treated malaria in the previous two weeks, suggesting the current infections were recrudescent parasitaemia or newly acquired infections. Interestingly, 2.0% of the positive cases were co-infected with SARS-CoV-2 (Figure 1C); They were children with no previous history of COVID-19 or malaria in the previous two weeks.

## Discussion and conclusion

Routine malaria surveillance in endemic communities is crucial to inform and assess the effectiveness of control interventions including diagnosis and antimalarial treatment. These interventions were considerably disrupted in many parts of Ghana as a result of the COVID-19 epidemics. Therefore, this study evaluated the malaria burden and its epidemiological overlaps with COVID-19 in southern Ghana.

Our findings in the Greater Accra and Central regions in October-December 2022, based on the self-reports, suggested that malaria was moderately prevalent, and varied heterogeneously, from 13% in the community setting to 27% in hospital settings. Among the reported cases, ∼71% reported using clinically prescribed antimalarials, likely representing high treatment-seeking in healthcare facilities; Indeed, we observed that participants enrolled in a hospital/clinic setting were up to 4 times more likely to report having malaria in the past two weeks compared to those enrolled in the community, i.e., public places. It is unlikely that this treatment-seeking was due to COVID-19 since the majority, 91.6%, had no history of COVID-19 in the last one month. Outpatient data collected during the COVID-19 epidemics in Ghana showed that treatment-seeking due to malaria increased in communities in the south ^8^ compared to the north ^3^. This uptick in treatment-seeking happened post-lockdowns and following rollout of COVID-19 vaccinations in early 2022.

Malaria parasite prevalence in the study communities in the Central region was 6%, comparable to the 6.8% prevalence (range 4-11%) that we estimated from a study of afebrile malaria cases in four communities located in the same region ^14^. All the detected cases and the majority of the COVID-19 positives, were asymptomatic and mostly in healthy adults, which is not unusual in high transmission settings like Ghana ^15^. Our data suggested that these asymptomatic malaria infections were recent infections or chronic parasitaemia, but we cannot rule out the possibility that they were recrudescent infections, particularly, among the RDT-positive cases with a history of clinically treated malaria in the past two weeks. Although clinical resistance to artemisinin combination therapies (ACTs), the standard treatment for uncomplicated malaria, has not been reported in Ghana ^16^, our study highlights the need to monitor and prioritise asymptomatic malaria infections during interventions.

By integrating malaria surveillance into COVID-19 programs, we uncovered epidemiological overlaps between the two diseases in our study communities. Particularly, the majority of the infections were asymptomatic, and mostly in the adults, with 2% co-infections. Our sample size for the RDT positive malaria cases was insufficient to detect any statistically significant association between *P. falciparum* and SARS-CoV-2 infections. Therefore, further investigations with larger sample sets are needed to evaluate the interplay between both infections within the host, and across populations.

In summary, the association between malaria and asymptomatic COVID-19 within the hospital settings underscores the need for accurate case diagnosis to inform effective clinical treatments. The CHEMS approach allowed us, researchers and clinicians, to effectively engage with the community, maximising the limited resources at our disposal to conduct co-surveillance activities for COVID-19 and malaria in southern Ghana.

## Data Availability

This study was embedded within a pre-clinical study for COVID-19 diagnostics test (Trial Registry # ACTRN12623000066684).

## Author Contributions

Conceptualisation: CAN. Funding acquisition: CAN & LM. Investigation and methods: All authors. Data curation: CAN, GA & MEA. Formal analysis: CAN, GA & MEA. Supervision: CAN, LM & GAA. Manuscript draft and formatting: CAN, GA & MEA. Review and editing: CAN, LM, GA, GAA, SA. Results visualisation/presentation: CAN & MEA. All authors have read and agreed to the published version of the manuscript.

## Institutional Review Board Statement

The study was reviewed and approved by the Institutional Review Boards of the Council for Scientific and Industrial Research (CSIR/IRB/AL/VOLl-023), Ghana Health Service (GHS-ERC 005/06/20), Basic and Applied Sciences, University of Ghana (ECBAS 063/19-20) and the University of Ghana Medical Centre (UGMC-MSRD).

## Informed Consent Statement

Study participants provided informed consent. Children under 18 years provided assent and their parents/legal guardian provided informed written consent prior to study enrolment.

## Acknowledgments

The CHEMS outreach programs were conducted with support from WACBIP, University of Ghana, with thanks to Francis Dzabeng (REDCap and data administrator) and the research assistants/technicians who assisted with enrolments and COVID-19 RT-PCR training. We thank the hospital staff, disease control officers, study site coordinators and the community leaders who contributed to CHEMS outreaches in the study communities. The entire study was largely successful because the study participants actively engaged in all the CHEMS activities, and voluntarily provided respiratory samples and demographic data.

